# mRNA-based COVID-19 booster vaccination is highly effective and cost-effective in Australia

**DOI:** 10.1101/2022.05.08.22274797

**Authors:** Rui Li, Hanting Liu, Christopher K Fairley, Jason J Ong, Yuming Guo, Zhuoru Zou, Li Xie, Guihua Zhuang, Yan Li, Mingwang Shen, Lei Zhang

**Author notes:** **Corresponding authors:** Lei Zhang, PhD, China-Australia Joint Research Center for Infectious Diseases, School of Public Health, Xi’an Jiaotong University Health Science Center, Xi’an, Shaanxi, China., Mingwang Shen, PhD, China-Australia Joint Research Center for Infectious Diseases, School of Public Health, Xi’an Jiaotong University Health Science Center, Xi’an, Shaanxi, China., Yan Li, PhD, Department of Population Health Science and Policy, Icahn School of Medicine at Mount Sinai, New York, NY, USA.

## Abstract

**Background:** Australia implemented an mRNA-based booster vaccination strategy against the COVID-19 Omicron variant in November 2021. We aimed to evaluate the effectiveness and cost-effectiveness of the booster strategy over 180 days.

**Methods:** We developed a decision-analytic Markov model of COVID-19 to evaluate the cost-effectiveness of a booster strategy (administered 3 months after 2^nd^ dose) in those aged ≥16 years in Australia from a healthcare system perspective. The willingness-to-pay threshold was chosen as A$ 50,000.

**Findings:** Compared with 2-doses of COVID-19 vaccines without a booster, Australia’s booster strategy would incur an additional cost of A$0.88 billion but save A$1.28 billion in direct medical cost and gain 670 quality-adjusted life years (QALYs) in 180 days of its implementation. This suggested the booster strategy is cost-saving, corresponding to a benefit-cost ratio of 1.45 and a net monetary benefit of A$0.43 billion. The strategy would prevent 1.32 million new infections, 65,170 hospitalisations, 6,927 ICU admissions and 1,348 deaths from COVID-19 in 180 days. Further, a universal booster strategy of having all individuals vaccinated with the booster shot immediately once their eligibility is met would have resulted in a gain of 1,599 QALYs, a net monetary benefit of A$1.46 billion and a benefit-cost ratio of 1.95 in 180 days.

**Interpretation:** The COVID-19 booster strategy implemented in Australia is likely to be effective and cost-effective for the Omicron epidemic. Universal booster vaccination would have further improved its effectiveness and cost-effectiveness.

**Funding:** National Natural Science Foundation of China. Bill and Melinda Gates Foundation

## Introduction

As of 5^th^ May 2022, the COVID-19 pandemic has caused more than 516 million cases and claimed more than 6 million lives worldwide^1^. Prevention and control of the disease have substantially interrupted economic activities and caused an enormous economic burden in both developing and developed countries^2-4^. Australia implemented a strict ‘COVID-zero’ policy and reported 200,000 cumulative infections and less than 2000 deaths (case-fatality rate ∼1%) over the past two years before the recent emergence of the Omicron variant^5^. However, the Omicron outbreak alone has caused more than 6 million new COVID-19 cases in six months since November 2021, despite more than 90% full (2-doses) vaccination coverage in Australia^6^. Given the lower viral pathogenicity of Omicron and high vaccination protection against disease progression, about 5000 deaths (case-fatality rate ∼0.08%) were reported for Omicron, demonstrating a much lower mortality rate compared with the previous variants.

A COVID-19 booster reportedly improves vaccine protection in fully vaccinated individuals against the Delta variant ^7-10^. Since the emergence of the Omicron variant, it has become apparent that the waning of vaccine protection of two doses of COVID-19 vaccine accelerated, likely due to Omicron’s ability to evade both natural and vaccine-induced immunity^11,12^. Australian Technical Advisory Group on Immunisation (ATAGI) approved the use of a booster first for individuals at greater risk of severe COVID-19 (e.g. immunocompromised individuals) at the end of October 2021 and gradually expanded to cover all individuals aged ≥16 years during the Omicron epidemic. ATAGI mainly recommends mRNA vaccines (Pfizer/BNT162b2, Moderna/mRNA1273) as a single booster dose over AstraZeneca for all eligible Australian residents, including those who have received the AstraZeneca COVID-19 vaccine for their primary course^13^. Among individuals who are fully vaccinated in Australia, about 35% were fully vaccinated with AstraZeneca and 65% with mRNA vaccines. For booster shots, mRNA vaccines have become the dominant booster option (>99%)^6^, although other vaccines also exist.

Accumulating evidence suggests that the disease severity associated with the Omicron variant is far lower than that of the Delta variant^14,15^, potentially contributing to much lower disease and economic burden. Most infections are asymptomatic or with very mild symptoms and often spontaneously recover within seven days, especially among fully vaccinated individuals^16,17^. An increasing number of experts argue for treating Omicron as a mild infectious disease like seasonal influenza in both surveillance and treatment^18,19^. Namely, non-pharmaceutical interventions (NPIs) for COVID-19 in Australia, similar to many other countries worldwide, would be minimal. As a result, vaccination becomes the major means to reduce COVID-19 transmission at a population level^5^. Therefore, evaluating the potential population impact and cost-effectiveness of the widely administered booster shots in Australia would shed important light on Australia’s COVID-19 prevention and control strategy. Our study aimed to evaluate the population impact and cost-effectiveness of Australia’s COVID-19 booster strategy during the Omicron epidemic using a decision-analytic Markov model.

## Methods

### Study design

Based on a decision-analytic Markov model, we conducted an economic evaluation on the cost-effectiveness of COVID-19 booster vaccination (predominately mRNA vaccines, 3 months after 2^nd^ dose) in those aged ≥16 years in Australia. The evaluation was conducted from a healthcare system perspective. The model was constructed using TreeAge Pro 2021 R1.1, and the analysis was conducted according to the Consolidated Health Economic Evaluation Reporting Standards 2022 (CHEERS 2022) statement^20^.

### Modelling

A decision-analytic Markov model was constructed to simulate the disease progression of SARS-CoV-2 infection for those aged ≥16 years over a period of 180 days. As the Omicron variant of SARS-COV-2 has become dominant in Australia, our study mainly focused on modelling the transmission of the Omicron variant. Existing evidence indicated that the vaccine efficacy (VE) of Pfizer-BioNTech BNT162b2, Moderna mRNA1273 and AstraZeneca AZD1222 against Omicron would gradually wane after three months^21-23^. Thus, we defined the vaccine efficacy from 2 weeks to 3 months after the 2^nd^ dose as a ‘short-term VE’, whereas the vaccine efficacy 3 months beyond the 2^nd^ dose as a ‘long-term VE’. Likewise, the vaccine efficacy after a booster could also be divided into a ‘short-term booster VE’ and a ‘long-term booster VE’ by the threshold of 3 months.

The model consisted of 11 health states capturing the disease progression of COVID-19 (Figure S1). A vaccinated individual may be infected by SARS-CoV-2 and enter a ‘latent infection state’. After a mean incubation period of 5.2 (4.1-7.0) days^24^, about 83% of infected individuals developed symptoms^25^, and the remaining asymptomatic infections would spontaneously recover. A symptomatic infection might first exhibit ‘mild/moderate’ symptoms. They might then ‘recover’ or deteriorate to a ‘severe’ state. A patient in the ‘severe’ state might ‘recover’ or progress to the ‘critical’ state. Similarly, a patient in the ‘critical’ state might ‘recover’ or ‘die’. Transition probabilities between states were estimated using the formula *p* = 1 − *e*^−*r*^, where r denoted daily transition rate^25^. The basic model cycle length was 1 day, with a half-cycle correction applied.

### Definition of scenarios

We explored three scenarios: Scenario 1: this counterfactual scenario assumed there was no booster vaccination implemented in Australia after the 2^nd^ dose; Scenario 2: current scenario represented the actual situation of booster vaccination in Australia, achieving coverage of 69.4% by 5^th^ May 2022; and Scenario 3: an ideal scenario of universal booster vaccination where all eligible individuals received a booster immediately once their booster vaccination requirement is met.

### Data collection

We collected data on the vaccine efficacy (VE) for Omicron variant infection based on an ongoing systematic review conducted by The International Vaccine Access Center^26^. We included 11 eligible studies to estimate the pooled short-term and long-term VE of the 2-dose vaccination and booster shot (Appendix 1.2). Based on the varied VE for preventing COVID-19 infection and severe progression, we developed a mathematical model to estimate the distributions of clinical disease stages after being infected by SARS-CoV-2 in vaccinated individuals compared to that in unvaccinated ones (Appendix 1.3). In addition, we collected Australia’s daily reported population incidence from ‘Our World in Data’, a website hosted and updated daily for COVID-19 cases worldwide^27^. Integrating with the COVID-19 vaccination information in Australia^6^, we calculated varied population incidence in varied vaccination statuses during the Omicron epidemic (Appendix 1.4).

The costs of booster vaccination included the costs of the mRNA-based vaccine in Australia (A$53/dose) and the vaccination administration ($20/dose)^28,29^. The cost of PCR tests and rapid antigen self-test for COVID-19 infection was estimated to be A$85 and A$13 per person, respectively^30,31^. In addition, we collected the costs of general practitioner (GP) consultation, general hospitalisation and ICU admission from published literature^32-34^. The cost of medical services varied across clinical disease stages^25,35-37^, and we calculated the total direct medical cost of COVID-19 cases with varied severity by multiplying the unit cost of the medical services by the duration of each disease stage (Appendix 1.5).Health utility scores for COVID-19 patients were derived from the disutility weights of severe lower respiratory tract infection^38,39^ and the estimates of pricing models for COVID-19 treatments published by the Institute for Clinical and Economic Review^40^ (Appendix 1.6).

We assumed a discount rate of 3% (0-6%) annually for both cost and quality-adjusted life-years (QALYs). We calculated the incremental costs and incremental QALYs for booster vaccination strategy compared with no booster (counterfactual scenario). The incremental cost-effectiveness ratio (ICER) was defined as the incremental cost per QALY gained. We used a willingness-to-pay (WTP) threshold of ICER<A$50,000. We conducted additional economic evaluations by calculating the benefit-cost ratio, net monetary benefit and cost/death saved.

### Sensitivity analysis

We conducted a univariate sensitivity analysis to examine the impact of model parameters within their respective ranges on the ICER to identify the most sensitive parameters and visualised the results using tornado diagrams (Figure 2). In addition, we conducted a probabilistic sensitivity analysis (PSA) based on 100,000 simulations to determine the probability of the booster strategy being cost-effective across a range of cost-effectiveness thresholds. The distributions of all model parameters are provided in Appendix 1.7. We also varied the evaluation period from 90 to 180 days and reported the corresponding ICER and benefit-cost ratio (Figure 3).

**Figure 1.**
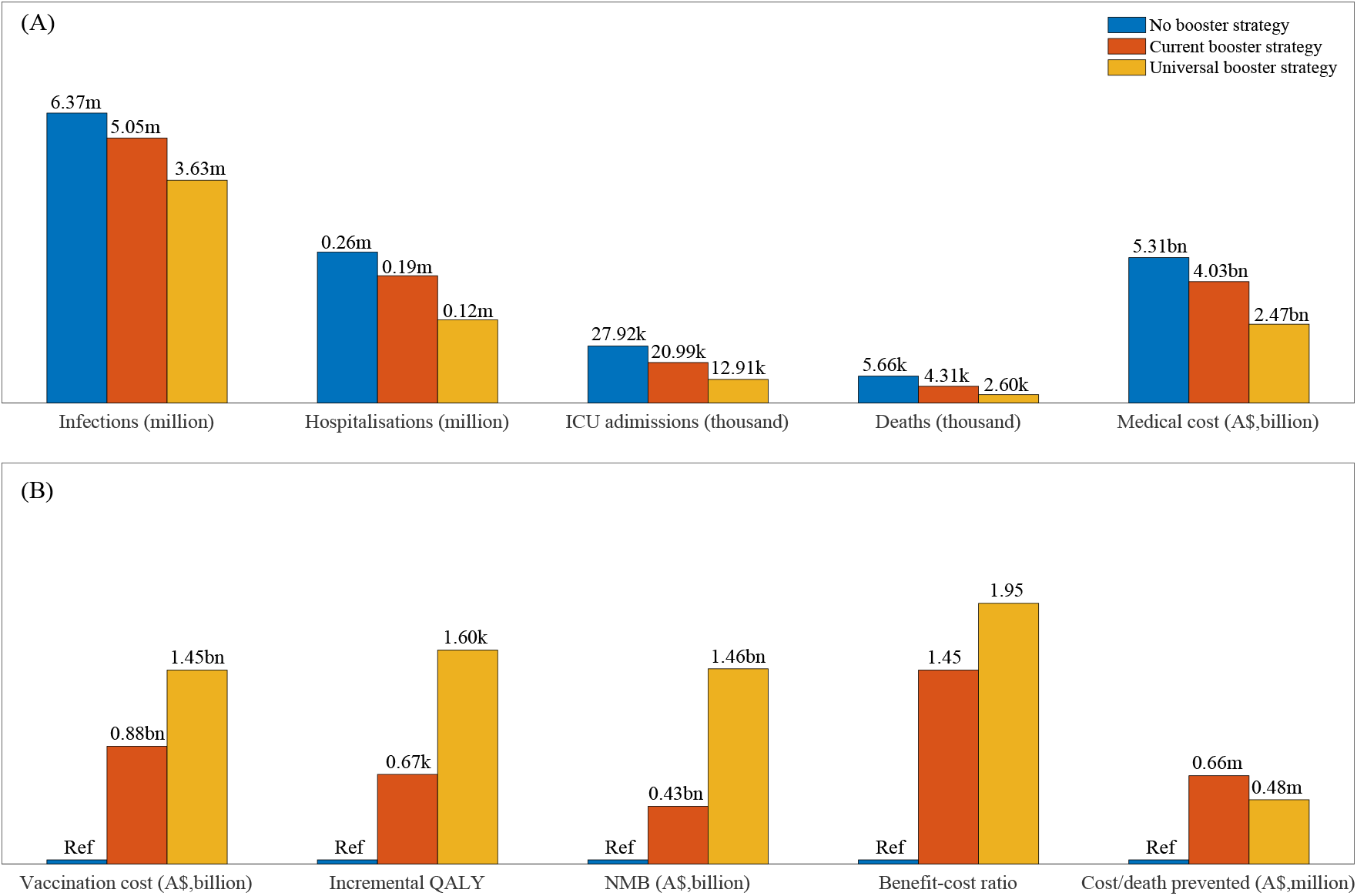
Comparison of population impacts and cost-effectiveness of the three booster vaccination scenarios for the Omicron epidemic in Australia. (No booster strategy: counterfactual scenario assumed there was no booster vaccination implemented in Australia after the 2^nd^ dose; Current booster strategy: current scenario represented the actual situation of booster vaccination in Australia; Universal booster strategy: universal scenario means all eligible individuals received a booster immediately once their booster vaccination requirement is met).

**Figure 2.**
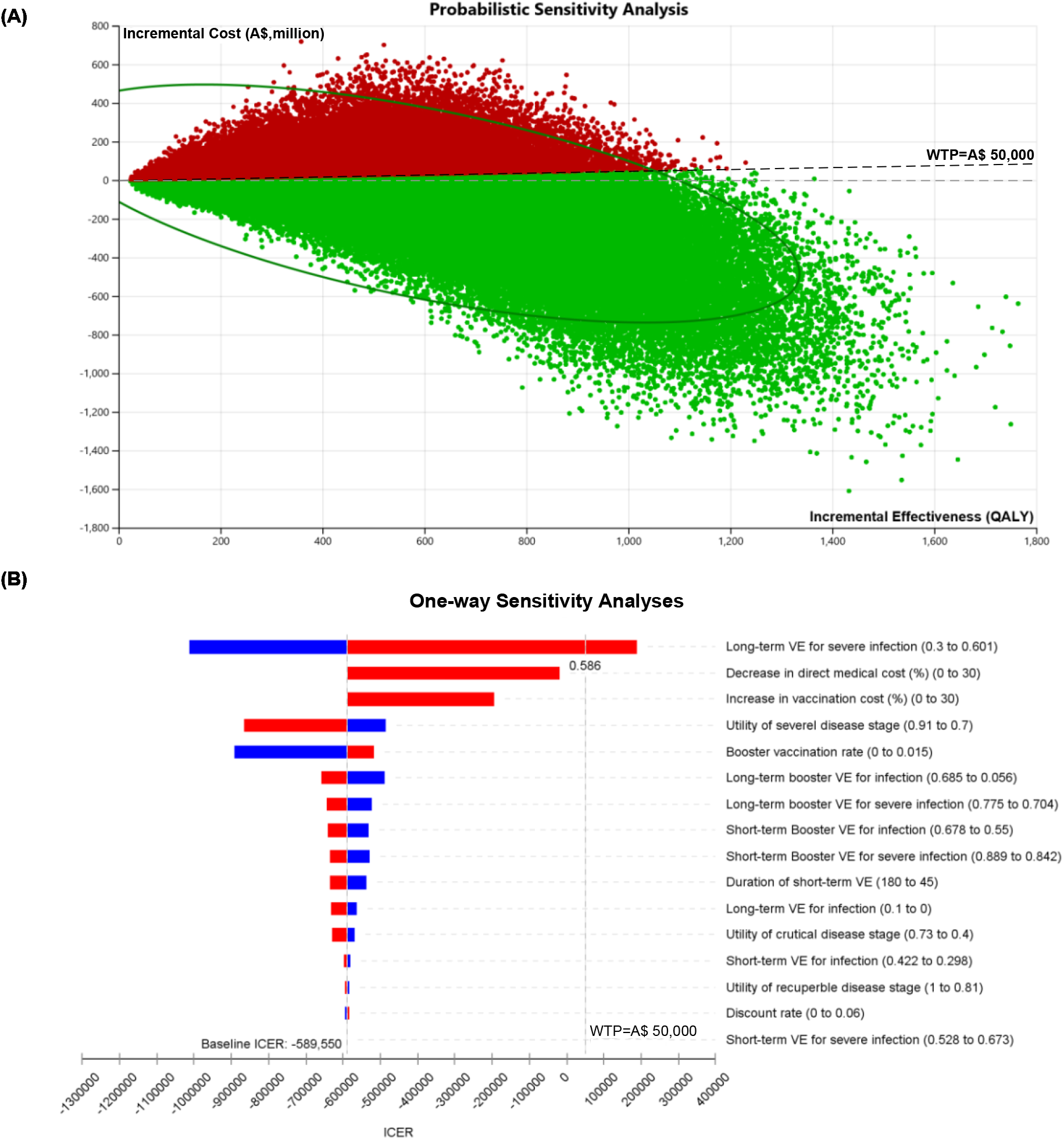
The cost-effectiveness analysis of COVID-19 booster vaccination strategy in Australia. (A) the result of probabilistic sensitivity analysis based on 100,000 simulations (71.2% chance of being cost-effective, including 66.7% chance of being cost-saving); (B) tornado plot of one-way sensitivity analyses. A horizontal bar was generated for each parameter analysis. The width of the bar indicates the potential effect of the associated parameter on the ICER when the parameter is changed within its range. The red part of each bar indicates high values of input parameter ranges, while the blue part indicates low values. The dotted vertical line represents the threshold of willingness-to-pay (WTP) of the baseline.

**Figure 3.**
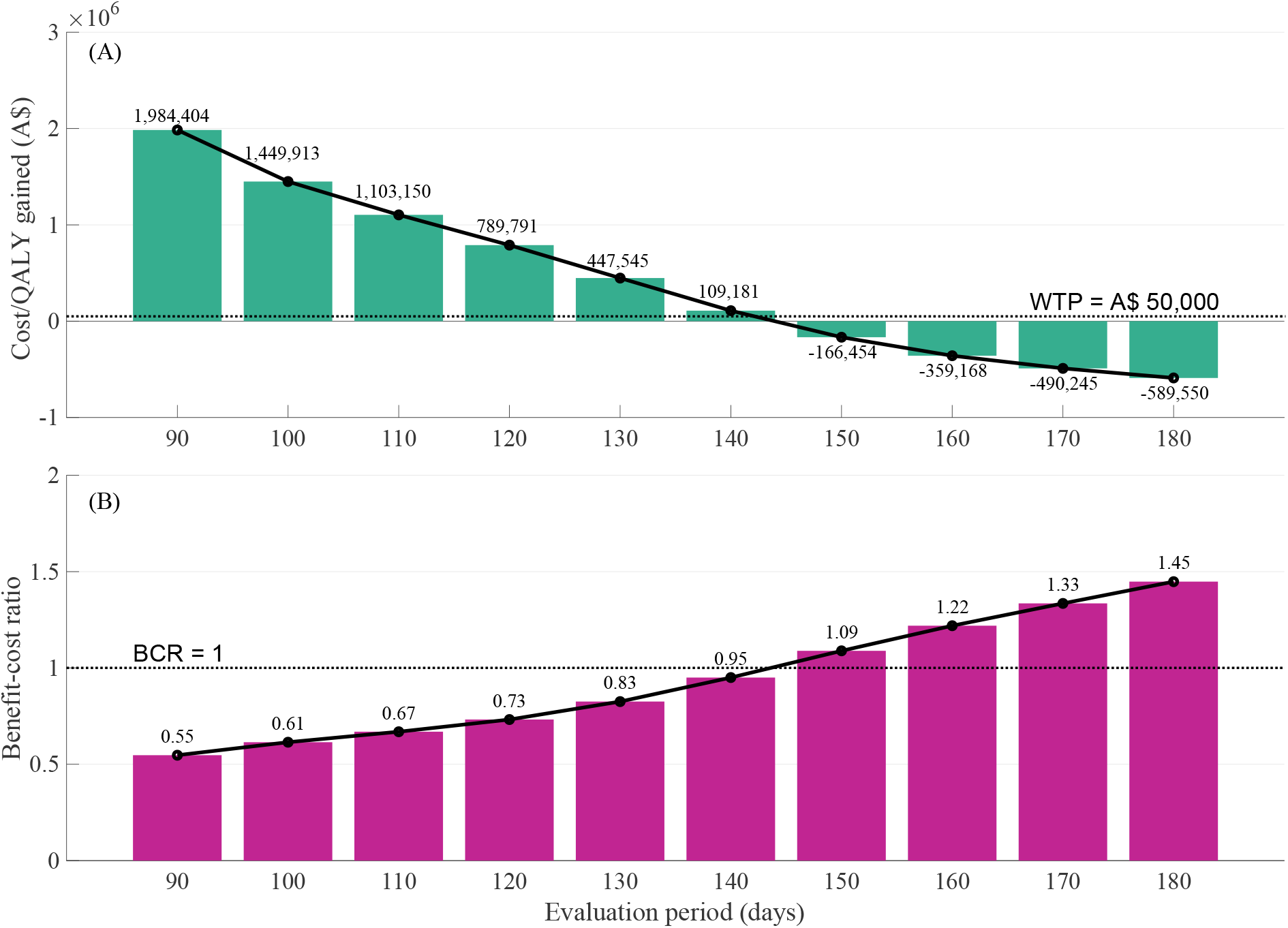
The effects of evaluation periods on the cost-effectiveness of the current booster strategy compared with the counterfactual ‘no booster strategy’. (A) Cost/QALY gained: incremental cost-effectiveness ratio; (B) Benefit-cost ratio, BCR.

## Results

### Population impact and cost-effectiveness of COVID-19 booster strategies in Australia

#### Counterfactual scenario with no booster strategy

Figure 1 compared various vaccination scenarios in Australia. In the counterfactual scenario where there was no booster vaccination in Australia, the Omicron epidemic would result in 6.37 million individuals being infected, 259,200 hospitalisations, 27,917 ICU admissions, and 5,656 deaths over an evaluation period of 180 days. This will amount to a direct medical cost of A$5.31 billion.

#### Current booster strategy

In comparison with the counterfactual scenario, the current booster strategy in Australia would lead to 1.32 million fewer infected cases, 65,170 fewer hospitalisations, 6,927 fewer ICU admissions and 1,348 fewer deaths in 180 days. At the same time, it would reduce the direct medical cost by A$1.28 billion but only incur an additional vaccination cost of A$0.88 billion, corresponding to a benefit-cost ratio of 1.45. We found that $655,077 was required to prevent one COVID-19 death. Moreover, the strategy would gain 670 QALYs during the 180 days with a net monetary benefit of A$0.43 billion. This suggested the current booster strategy is cost-saving.

#### Universal booster strategy

Compared with the current scenario, a universal booster strategy, where all eligible individuals receive a booster immediately when their booster vaccination requirement is met, would further improve the benefits. In addition to the gains in the current booster strategy, this scenario would further reduce 1.42 million infected cases, 73,753 hospitalisations, 8,081 ICU admissions and 1,706 deaths, which amount to A$1.56 billion less in direct medical costs. Likewise, the universal booster strategy would gain 1,599 QALYs and a net monetary benefit of A$1.46 billion during the 180 days with a benefit-cost ratio of 1.95. This scenario would incur an additional vaccination cost of A$1.45 billion and cost A$476,123 to prevent one COVID-19 death.

### Impact of individual model parameters on booster cost-effectiveness

The PSA, based on 100,000 simulations, demonstrated the probability of being cost-effective (including being cost-saving) with the current booster strategy was 71.2%, indicating a high chance of being cost-effective compared with no booster vaccination (**Figure 2A**). In contrast, the tornado diagram demonstrated that varying any individual model parameter except long-term VE for severe infection at one time would not change the conclusion of cost-effectiveness of the booster strategy (**Figure 2B**). We also noted that both the increase in vaccination cost and decrease in direct medical cost for COVID-19 treatment would reduce the cost-effectiveness of the booster strategy but were not sufficient to alter our conclusion.

### Impact of varied evaluation periods on booster cost-effectiveness

Figure 3 investigated the impact of varying evaluation periods (from 90 to 180 days) on the cost-effectiveness of the booster strategy compared to no booster. We found that for the current booster strategy to be cost-effective (ICER<A$50,000), the evaluation period in this cost-effectiveness analysis needed to be >140 days (**Figure 3A**). This indicated that the booster strategy would be more cost-effective and beneficial in the long term than in the short term. Our results showed that the benefit-cost ratio of a booster strategy was 0.55 in a 90-day evaluation period, which meant that every dollar of investment spent on the booster would only save 0.55 dollars for treating fewer hospitalised COVID-19 patients. However, this ratio would increase from 0.55 to 1.45 when the evaluation period was extended to 180 days (**Figure 3B**).

## Discussion

Our study evaluated the cost-effectiveness of the mRNA-based booster vaccination strategy among individuals aged ≥16 years in Australia. We identified several key findings. First, the current booster vaccination strategy is highly cost-saving. Compared to the no booster scenario, the current booster strategy would prevent 1.32 million new infections, 65,170 hospitalisations, 6,927 ICU admissions and 1,348 deaths in 180 days. It demonstrates a cost-benefit ratio of 1.45 and a net monetary benefit of A$0.43 billion. Further, a universal vaccination strategy of having all individuals vaccinated with the booster shot immediately once their eligibility is met would be even more cost-effective compared with the current scenario, further preventing 1.42 million additional new cases, 73,753 hospitalisations, 8,081 ICU admissions and 1,706 deaths. It demonstrates a cost-benefit ratio of 1.95 and a net monetary benefit of A$1.46 billion.

Our findings of cost-effectiveness are comparable with the findings from similar studies in other settings^41,42^. A recent cost-effectiveness analysis demonstrates that a BNT162n booster strategy is cost-saving among individuals aged ≥65 years in the United States (US)^42^, with a benefit-cost ratio of 1.95. Compared to this 1.95 ratio, our study identifies a lower benefit-cost ratio of 1.45 in Australia, which is mostly attributed to the fact that vaccine price is twice the cost in the US^28,43^. A higher vaccine price due to export and transport would reduce the cost-effectiveness of a booster strategy but not enough to alter the conclusion. Further, the significant development of antiviral drugs for COVID-19 treatment holds promise to change the pandemic’s course^44^. Pfizer’s oral antiviral drug Paxlovid is highly effective and reduces by 89% the number of hospital admissions and deaths among people with COVID-19 who are at high risk of severe illness^45^. Similarly, molnupiravir and fluvoxamine have also shown high efficacy in preventing severe isllness^46^. However, supply shortages and huge global inequities in access to new treatments for COVID-19, especially in low- and middle-income countries^47,48^, may be major obstacles to the scale-up of these antiviral treatments worldwide in the short term. In Australia, the first oral treatments for COVID-19, such as molnupiravir and Paxlovid were provisionally approved by the Therapeutic Goods Administration (TGA) on 18^th^ January 2022. If low-price antiviral drugs can be made available and accessible to the general population like vaccines in the future, the cost-effectiveness of the booster strategy will likely reduce substantially.

Our study demonstrates a universal booster strategy would have further improved the strategy’s cost-effectiveness. This finding strongly suggests that rapid scale-up of vaccination would be the most beneficial strategy in combating the Omicron epidemic in Australia. The main reason for the excellent cost-effectiveness of the universal vaccination strategy is that the vaccine effectiveness (VE) of 2-doses against the Omicron variant is low, and it declines sharply over time^21-23^. Based on our meta-analysis (Appendix 1.2), the short-term VE against Omicron infections of 2 doses is 36% and would reduce to only 4% after a period of 90 days. In contrast, the short-term VE against Omicron infections of a booster shot is 62% and would reduce to 46% after 90 days. Hence, for the Omicron epidemic with an increased incidence of reinfections and breakthrough infections^49^, receiving a booster as early as possible may be the most cost-effective way to reduce the COVID-19 disease burden.

Our study suggests the vaccination booster strategy is practical for COVID-19 prevention and control in Australia. To date, Australia has fully opened its borders, removing most social-distancing restrictions (such as city lockdown) and the requirement of facial masks in most public areas, including indoors. Contact tracing and the quarantine requirements for close contacts have also been lifted in Australia. Living with the virus has become a socially accepted norm in the post-COVID era. Accumulating evidence indicates that the epidemic will become endemic. Our study suggests that the epidemic needs to be persisting for at least 140 days for the booster strategy to be cost-effective. This implies the booster strategy will not be compatible with the previous ‘COVID-zero’ strategy, which often reduced the epidemic to a low level within weeks through harsh NPIs but is more aligned with an endemic COVID-19 control in Australia. Also, considering the rapid emergence of new SARS-COV-2 mutants^50,51^, enhancing the vaccine protection in the population remains the most practical way to avoid a sudden surge of the disease burden of COVID-19 and potential overload of the healthcare system in Australia. Further, learning from the experiences of seasonal influenza control^52^ and regular improvement of COVID-19 vaccines may also be a necessary strategy in the long term.

Our study has several limitations. First, in the absence of empirical evidence of randomised controlled trials, we estimated the efficacy of the mRNA-based booster against the Omicron variant based on a synthesis of evidence from real-world data. We conducted various sensitivity analyses to account for the uncertainty in parameter estimation based on real-world data. Second, we assumed that the efficacy of COVID-19 vaccines begins to wane in 3 months. In reality, the efficacy of vaccines is more likely to gradually decline without a clear cut-off. This assumption may have led to an overestimate of vaccine efficacy in the short term, and an underestimate in the long term. Third, we did not consider the other non-mRNA-based COVID-19 vaccines (e.g., AstraZeneca or Novavax) in Australia. However, AstraZeneca is no longer recommended for use as the booster dose for people who received a primary vaccination course of the AstraZeneca COVID-19 vaccine, and Novavax is rarely used (<1%) in Australia. Finally, given limited public health resources and escalating health inequity during the pandemic, there is a need for more targeted, local-based vaccine and booster distribution strategies that can achieve a tradeoff between cost-effectiveness and equity. The design of such strategies, while beyond the scope of this work, will be critical in alleviating the burden of COVID-19, reducing health care costs, and achieving equity.

In conclusion, Australia’s current COVID-19 booster strategy is likely to be effective and cost-effective for curbing this Omicron epidemic. Further, achieving a universal booster vaccination would have improved its effectiveness and cost-effectiveness.

**Table 1.** The results of the cost-effectiveness analysis of COVID-19 booster vaccination in Australia over an evaluation period of 180 days.

|  | Counterfactual<br>scenario with<br>no booster<br>strategy | Current booster<br>strategy | Universal<br>booster<br>strategy | Incremental<br>benefits of<br>booster<br>strategy* | Incremental<br>benefits of<br>universal booster<br>strategy* |
| --- | --- | --- | --- | --- | --- |
| QALY | 10,082,957 | 10,083,628 | 10,084,557 | 670 | 1599 |
| Costs (A\$, million) | 5,311.5 | 4,841.3 | 3,928.8 | -395.2 | -1,382.7 |
| Booster vaccination cost | 0 | 883.0 | 1,454.3 | 883.0 | 1,454.3 |
| Direct medical cost | 5,311.5 | 4,033.3 | 2,474.5 | -1,278.2 | -2,837.0 |
| Death cases | 5,656 | 4,308 | 2,602 | 1,348 | 3,054 |
| ICER | -- | -- | -- | Cost-saving | Cost-saving |
| Benefit-cost ratio | -- | -- | -- | 1.45 | 1.95 |
| Cost/death prevented, A\$ | -- | -- | -- | 655,077 | 476,123 |
| Net monetary benefit, (A\$,<br>million) | -- | -- | -- | 428.7 | 1,462.7 |

\* Incremental benefits = difference in QALY while comparing to the no booster strategy.
Benefit-cost ratio: the ratio between the reduction in the direct medical cost for COVID-19 and the investment in booster vaccination for individuals aged $\geq 16$ in Australia.
Net monetary benefit (NMB) is calculated as (incremental benefit × willingness-to-pay threshold) – incremental cost.

## Data Availability

All data produced in the present work are contained in the manuscript

## Declarations

### Ethics approval and consent to participate

Not applicable

### Authors’ contributions

LZ conceived the study. LZ, YL, MS and RL designed and constructed the model. RL performed the modelled analyses, graphed and interpreted the results. RL, HL, ZZ, and LX contributed to the collection of data and model parameters. RL drafted the manuscript. LZ, MS, YL, CKF, JJO and YG critically revised the manuscript. All authors reviewed the manuscript and approved the final version.

