## Appendix for "mRNA-based COVID-19 booster vaccination is highly effective and cost-effective in Australia"

This supplementary document describes in detail the model construction and estimation of parameters presented in the main text.

**1.1 Health stages**

We expanded the previous published decision-analytic Markov model to simulate the disease progression of SARS-CoV-2 infection in a designated initial cohort of 100,000 individuals over a period of 180 days. The model consisted of 10 health states depicting varied disease progression of COVID-19 **(Figure S1).** Existing evidence indicated that the vaccine efficacy (VE) of Pfizer-BioNTech BNT162b2, Moderna mRNA1273 and ChAdOx1 nCoV-19 AZD1222 against Omicron would gradually wane after three months^1-3^. We defined the 11 diseases states as follows.

- Short-term VE: the vaccine efficacy from 2 weeks to 3 months after the 2nd dose of vaccines
- Long-term VE: the vaccine efficacy 3 months beyond the 2nd dose
- Short-term booster VE: the vaccine efficacy from 2 weeks to 3 months after the 2nd dose of vaccines
- Long-term booster VE: the vaccine efficacy 3 months beyond the 2nd dose
- Incubation: cases prior to symptom onset
- Asymptomatic: cases who never developed symptoms ever throughout the course of their disease
- Mild/moderate: cases without pneumonia and cases with mild pneumonia
- Severe: cases who developed dyspnoea and/or hypoxemia and managed in a hospital but not requiring intensive care unit (ICU)
- Critical: cases who developed respiratory failure, and/or septic shock, and/or multiple organ dysfunction/failure and managed in an ICU; some of them who recuperated from critical disease need to go through the recuperation stage-remaining in the hospital or other health care facility
- Recovered: cases who recovered from infection stages; we assumed that a recovered individual could not be reinfected for the next 180 days (similar to the short-term vaccine protection)
- Death: COVID-19 related death

A vaccinated individual may be infected by SARS-CoV-2 strains and suffered a COVID-19 disease with varied progression as follows.

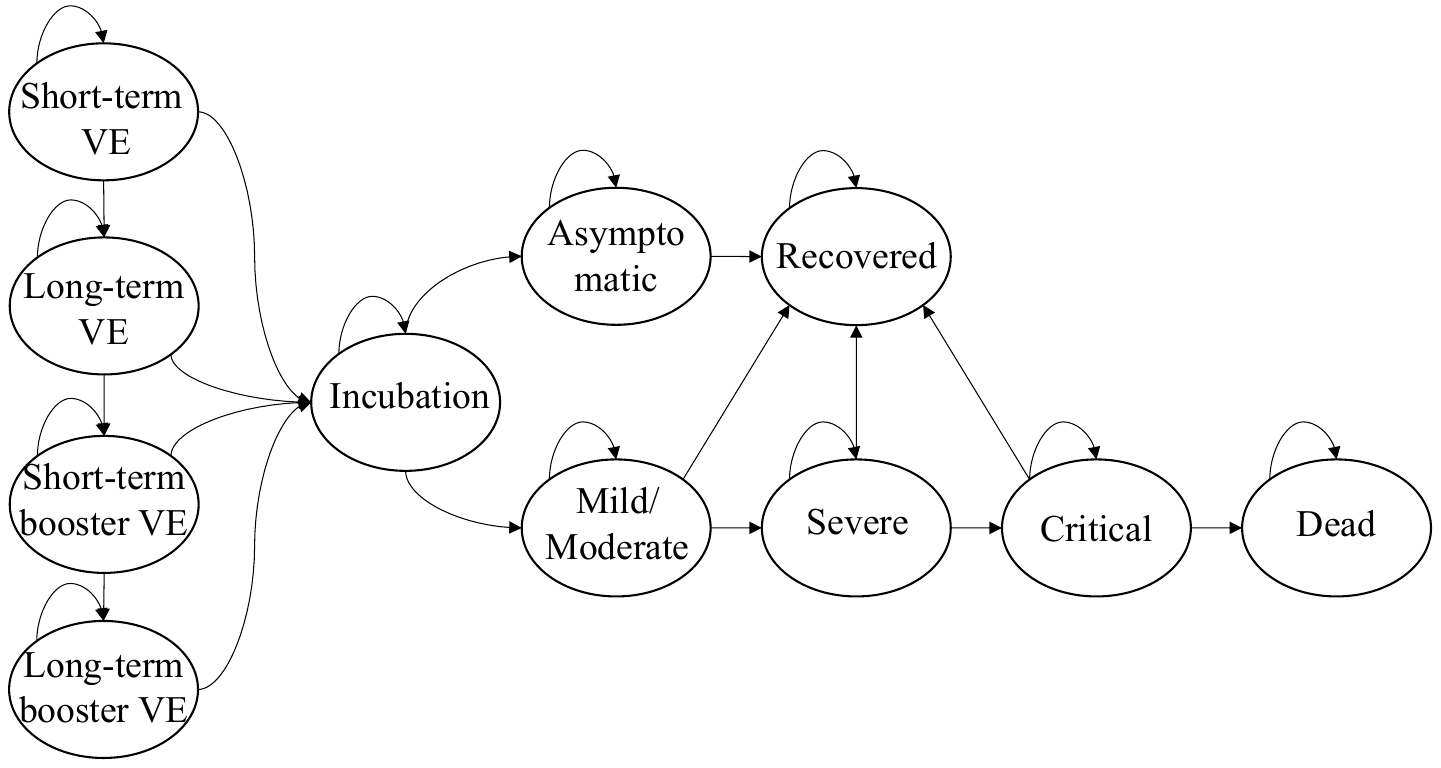

**Figure S1.** Markov model of COVID-19 natural history in the vaccinated population

**1.2 Vaccine efficacy against Omicron variant**

We collected information on the vaccine efficacy (VE) for Omicron variant infection in all ages based on an ongoing systematic review conducted by The International Vaccine Access Center^4^. After elimination of irrelevant literature, we finally included eleven studies from the review and extracted the original incident data to perform our own meta-analysis. We contacted authors of eligible studies to request additional information when the required data could not be extracted from the paper.

Overall, eleven studies were included in quantitative data synthesis and all of them were designed by test-negative case-control studies (Odds ratio, OR, as risk measurement). The studies aimed to compare the risk of infection and progressing severe COVID-19 disease against Omicron between individuals who were fully vaccinated or got a booster shot by BNT162b2, mRNA1273 or AZD1222 (**Table S2-3**). According to the timing after getting a second or booster shot (90 days), we artificially divided the VE into different period due to significant difference of VE^1,3^ (**Figure S2**).

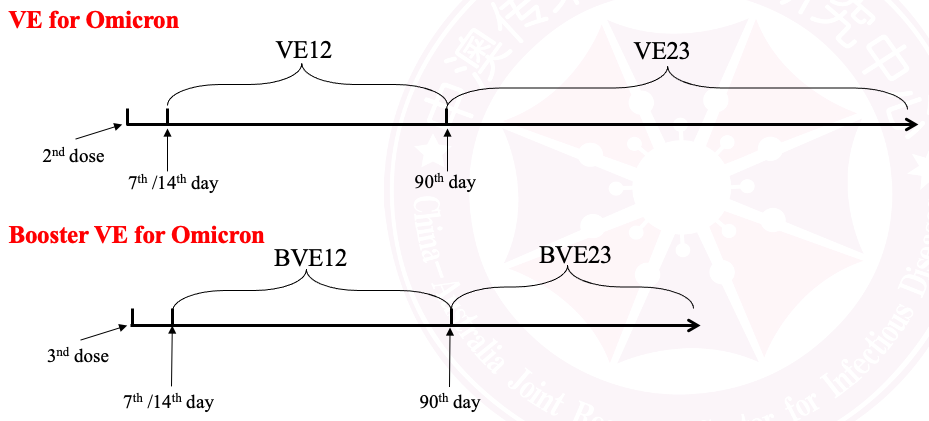

**Figure S2**. Waning vaccine effectiveness with time against Omicron in individuals who were fully vaccinated or got a booster shot.

We extracted the original incidence data form included studies and several meta-analyses to summarise the pooled OR, grouped by vaccination period and outcome measure. We then calculated the vaccine efficacy using the formula: (1–OR) multiplied by 100% as follow.

| **Variables** | **Preventing infection for Omicron** | | **Preventing severe infection for Omicron** | |
| --- | --- | --- | --- | --- |
|  | Pooled OR | VE | Pooled OR | VE |
| VE12 | 0.637 (0.578, 0.702) | **36.3% (29.8, 42.2)** | 0.393 (0.327, 0.472) | **60.7% (52.8, 67.3)** |
| VE23 | 0.963 (0.900, 1.030) | **3.7% (-3.0, 10.0)** | 0.528 (0.399, 0.700) | **47.2% (30.0, 60.1)** |
| BVE12 | 0.381 (0.322, 0.450) | **61.9% (55.0, 67.8)** | 0.132 (0.111, 0.158) | **86.8% (84.2, 88.9)** |
| BVE23 | 0.545 (0.315, 0.944) | **45.5% (5.6, 68.5)** | 0.258 (0.225, 0.296) | **74.2% (70.4, 77.5)** |

**1.3 Distribution of clinical disease stages of various vaccination status**

We collected the distribution of clinical disease stages after being infected by Omicron variant in fully vaccinated group from COVID-19 Australia Epidemiology Report and calibrated by published literature^5,6^. Based on the varied VE of short-term, long-term and booster, we developed a mathematical model to estimate the similar distributions in unvaccinated and other vaccinated group (**Figure S3**). In the model, we calculated the numerator and denominator of the proportion of severe infections in vaccinated patients ($P_{vs}$) by three factors, which are the proportion of severe infections in unvaccinated infections ($P_{s}$), the vaccine efficacy for preventing a SARS-COV-2 infection ($V_{i}$) and for preventing a severe COVID-19 case ($V_{s}$).

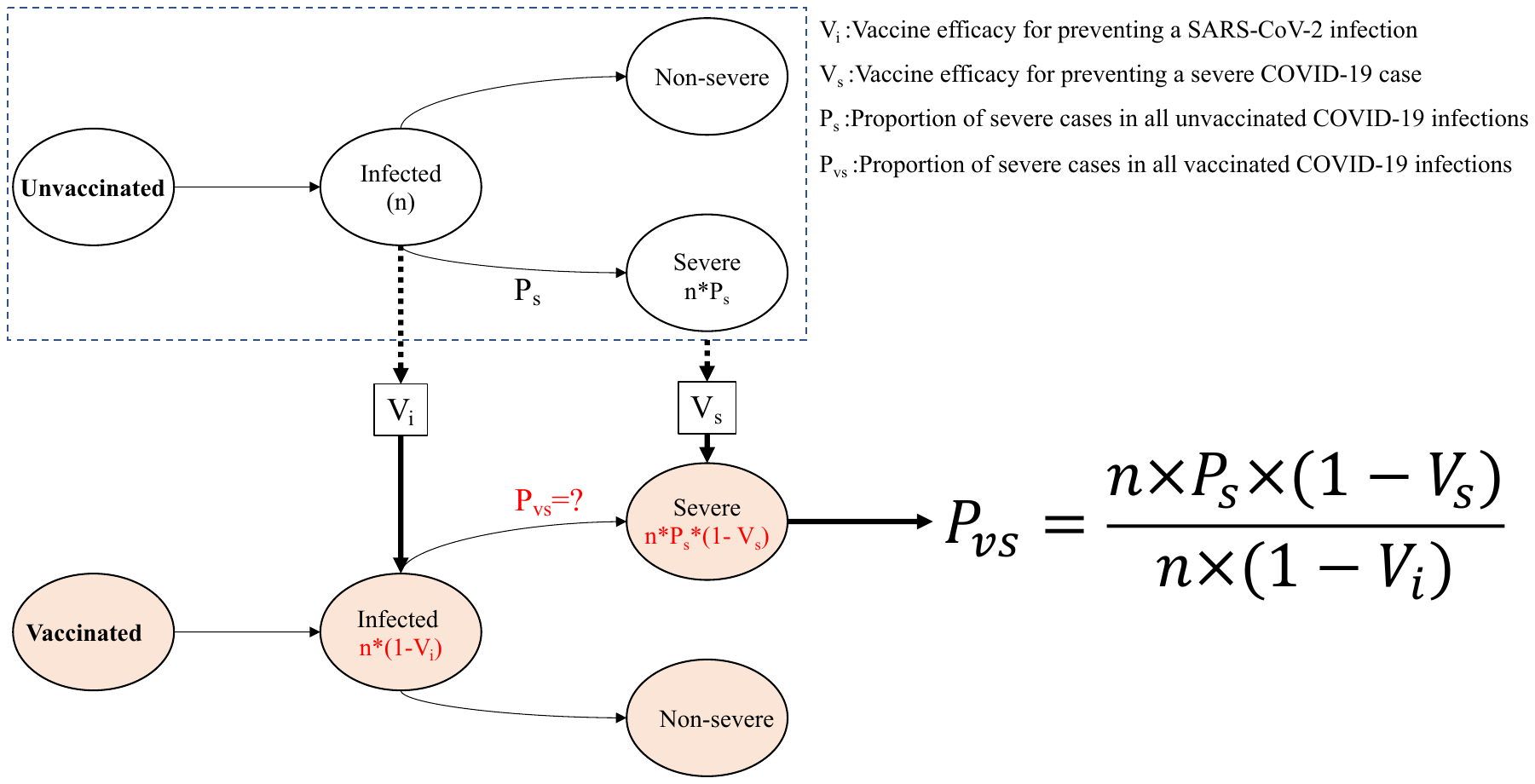

**Figure S3**. The model flowchart to estimate the proportion of severe cases in all COVID-19 infections.

Then, we could obtain the proportion of the severe cases (including severe and critical) after COVID-19 infection in various vaccination status. We assumed the proportion of critical cases in severe infections was fixed both in unvaccinated and vaccinated groups. Thus, we could calculate the proportion of severe and critical clinical outcome after COVID-19 infections in varied vaccinated group, according to that in unvaccinated group. Similarly, we calculated the proportion of asymptomatic and mild/moderate clinical outcome after COVID-19 infections in varied vaccinated group. The details of the distribution are list as follow.

| Clinical outcomes | The distribution in unvaccinated group | The distribution of long-term VE group | The distribution of short-term VE group | The distribution of short-term booster VE group | The distribution of long-term booster VE group |
| --- | --- | --- | --- | --- | --- |
| Asymptomatic infection | 16.88% | 17.42% | 17.52% | 17.80% | 17.62% |
| Mild/Moderate illness | 75.43% | 77.84% | 78.27% | 79.54% | 78.74% |
| Severe illness | 6.83% | 4.21% | 3.74% | 2.37% | 3.23% |
| Critical illness | 0.85% | 0.53% | 0.47% | 0.30% | 0.40% |
| Critical (recover) | 0.66% | 0.41% | 0.36% | 0.23% | 0.31% |
| Critical (die) | 0.19% | 0.12% | 0.10% | 0.07% | 0.09% |

**1.4 Population incidence of COVID-19 Omicron strain.**

We collected the daily reported population incidence in Australia from Our World in Data, a website provided international COVID-19 epidemic dataset^7^. Combined the COVID-19 vaccination in Australia^8^, we calculated varied population incidence in varied vaccinated group among the Omicron epidemic.

**1.5 Direct medical cost**

Based on COVID-19 management of Medicare Benefits Schedule in Australia^9^, we collected the relevant items of direct medical cost due to COVID-19 infection as follow:

| Items | Cost | Resource |
| --- | --- | --- |
| Per PCR test for COVID-19 infection, A$ | 85 | ^10^ |
| Per rapid antigen self-test for COVID-19 infection, A$ | 13.25 | ^11^ |
| Per general practitioner (GP) consultation, A$ | 37 | ^12^ |
| Per day of general hospitalization, A$ | 1300.9 | ^13^ |
| Per day of ICU hospitalization, A$ | 4822.3 | ^14^ |
| Per dose of COVID-19 vaccine booster, A$ | 53 | ^15^ |
| Per dose of vaccine administration, A$ | 19.95 | ^16^ |

Combined the coverage of different way of medical services and duration of varied clinical disease stages^5,17-19^, we calculated total direct medical cost of a COVID-19 cases with varied clinical severity outcomes.

| Clinical outcomes | Proportion of GP consultation | Proportion of hospitalisation | Duration of varied clinical disease stages (days) | Total cost, A$ |
| --- | --- | --- | --- | --- |
| Asymptomatic infection | 0% | 0% | 0 | 0 |
| Mild/Moderate illness | 100% | 0% | 10 | 37 |
| Severe illness | 100% | 100% | 6.5+10.5 | 13696.5 |
| Critical (recover) illness | 100% | 100% | 3+7.1+11.9 | 41046.4 |
| Critical (die) illness | 100% | 100% | 3+7.1+11.9+5.7 | 74073.4 |

**1.6 Health state utilities**

We collected utility scores for COVID-19 patients from the disutility weights of severe lower respiratory tract infection^20,21^ and the estimates of pricing models for COVID-19 treatments published by the Institute for Clinical and Economic Review^22^. We calculated the average utility from two sources and adopted their lowest and highest bounds^23^.

| Health states | Disability weight 1 | Disability weight 2 | **Average utility** |
| --- | --- | --- | --- |
| In asymptomatic state | — | — | **1** |
| In mild/moderate state | — | — | **1** |
| In severe state | 0.13 (0.09, 0.19) | 0.30 | **0.785 (0.700, 0.910)** |
| In critical state | 0.41 (0.27, 0.56) | 0.50-0.60 | **0.520 (0.400, 0.730)** |
| In recuperable state | — | 0.19 | **0.905 (0.810, 1.000)** |
| In dead state | 0 | 0 | **0** |

**1.7 Sensitivity analyses**

We extensively explored the impact of model parameters on the baseline results with univariate and probabilistic sensitivity analyses (PSA). Univariate sensitivity analysis is for each of 16 parameters and varying one parameter at one time in their range. PSA allows variation of all 16 parameters together at one time in their distribution.

| **No** | **Name** | **Distribution** | **Reference** |
| --- | --- | --- | --- |
| 1 | Duration of short-term VE | Uniform (45, 180) | Assumed and ^1-3^ |
| 2 | Short-term VE for preventing Omicron infection | Triangular (0.298, 0.363, 0.422) | Appendix 1.2 |
| 3 | Short-term VE for preventing severe COVID-19 disease | Triangular (0.528, 0.607, 0.67.3) | Appendix 1.2 |
| 4 | Long-term VE for preventing Omicron infection | Triangular (0, 0.037, 0.100) | Appendix 1.2 |
| 5 | Loge-term VE for preventing severe COVID-19 disease | Triangular (0.300, 0.472, 0.601) | Appendix 1.2 |
| 6 | Short-term booster VE for preventing infection, compared with no vaccinated | Triangular (0.550, 0.619, 0.619) | Appendix 1.2 |
| 7 | Short-term booster VE for preventing severe infection, compared with no vaccinated | Triangular (0.842, 0.868, 0.889) | Appendix 1.2 |
| 8 | Long-term booster VE for preventing infection, compared with no vaccinated | Triangular (0.056, 0.455, 0.685) | Appendix 1.2 |
| 9 | Long-term booster VE for preventing severe infection, compared with no vaccinated | Triangular (0.704, 0.742, 0.775) | Appendix 1.2 |
| 10 | Booster vaccination rate | Uniform (0, 0.015) | Assumed and ^8^ |
| 11 | Decrease in direct medical cost (%) | Uniform (0, 30) | Assumed |
| 12 | Increase in vaccination cost (%) | Uniform (0, 30) | Assumed |
| 13 | Utility of severe disease stage | Triangular (0.700, 0.785, 0.910) | Appendix 1.6 |
| 14 | Utility of critical disease stage | Triangular (0.400, 0.520, 0.730) | Appendix 1.6 |
| 15 | Utility of recuperable disease stage | Triangular (0.810, 0.905, 0.100) | Appendix 1.6 |
| 16 | Discount rate | Uniform (0.00, 0.06) | ^24^ |

3.	Hiam Chemaitelly HHA, Sawsan AlMukdad, Patrick Tang, Mohammad R. Hasan, Hadi M. Yassine, Hebah A. Al Khatib, Maria K. Smatti, Peter Coyle, Zaina Al Kanaani, Einas Al Kuwari, Andrew Jeremijenko, Anvar Hassan Kaleeckal, Ali Nizar Latif, Riyazuddin Mohammad Shaik, Hanan F. Abdul Rahim, Gheyath K. Nasrallah, Mohamed Ghaith Al Kuwari, Adeel A. Butt, Hamad Eid Al Romaihi, Mohamed H. Al-Thani, Abdullatif Al Khal, Roberto Bertollini, Laith J. Abu-Raddad. Duration of protection of BNT162b2 and mRNA-1273 COVID-19 vaccines against symptomatic SARS-CoV-2 Omicron infection in Qatar. 2022.

4.	IVAC. Results of COVID-19 Vaccine Effectiveness Studies: An Ongoing Systematic Review. 2022. <https://view-hub.org/resources> (accessed Mar 05, 2022).

5.	COVID-19 Australia: Epidemiology Report 57: Reporting period ending 16 January 2022. *Commun Dis Intell (2018)* 2022; **46**.

6.	Wang L, Berger NA, Kaelber DC, Davis PB, Volkow ND, Xu R. COVID infection rates, clinical outcomes, and racial/ethnic and gender disparities before and after Omicron emerged in the US. *medRxiv* 2022.

25.	nnabel A Powell FK, Julia Stowe, Kelsey McOwat, Vanessa Saliba, Mary E Ramsay, Jamie Lopez-Bernal, Nick Andrews, Shamez N Ladhani. Adolescent vaccination with BNT162b2 (Comirnaty, Pfizer-BioNTech) vaccine and effectiveness against COVID-19: national test-negative case-control study, England. 2022.

26.	Collie S, Champion J, Moultrie H, Bekker LG, Gray G. Effectiveness of BNT162b2 Vaccine against Omicron Variant in South Africa. *N Engl J Med* 2022; **386**(5): 494-6.

27.	Hung Fu Tseng BKA, Yi Luo, Lina S. Sy, Carla A. Talarico, Yun Tian, Katia J. Bruxvoort, Julia E. Tubert, Ana Florea, Jennifer H. Ku, Gina S. Lee, Soon Kyu Choi, Harpreet S. Takhar, Michael Aragones, Lei Qian. Effectiveness of mRNA-1273 against SARS-CoV-2 omicron and delta variants. 2022.

28.	Sarah A. Buchan HC, Kevin A. Brown, Peter C. Austin, Deshayne B. Fell, Jonathan B. Gubbay, Sharifa Nasreen, Kevin L. Schwartz, Maria E. Sundaram, Mina Tadrous, Kumanan Wilson, Sarah E. Wilson, Jeffrey C. Kwong. Effectiveness of COVID-19 vaccines against Omicron or Delta symptomatic infection and severe outcomes. 2022.

29.	Tartof SYaS, Jeff M. and Puzniak, Laura and Hong, Vennis and Xie, Fagen and Ackerson, Bradley K. and Valluri, Srinivas R. and Jodar, Luis and McLaughlin, John M. BNT162b2 (Pfizer–Biontech) mRNA COVID-19 Vaccine Against Omicron-Related Hospital and Emergency Department Admission in a Large US Health System: A Test-Negative Design. 2022.

30.	Adam S. Lauring MWT, James D. Chappell, Manjusha Gaglani, Adit A. Ginde, Tresa McNeal, Shekhar Ghamande, David J. Douin, H. Keipp Talbot, Jonathan D. Casey, Nicholas M. Mohr, Anne Zepeski, Nathan I. Shapiro, Kevin W. Gibbs, D. Clark Files,et al. Clinical Severity and mRNA Vaccine Effectiveness for Omicron, Delta, and Alpha SARS-CoV-2 Variants in the United States: A Prospective Observational Study.

31.	Katrina Spensley SG, Paul Martin, Tina Thomson, Candice L. Clarke, Graham Pickard, David Thomas, Stephen P. McAdoo, Paul Randell, Peter Kelleher, Rachna Bedi, Liz Lightstone, Maria Prendecki, Michelle Willicombe. Comparison of vaccine effectiveness against the Omicron (B.1.1.529) variant in patients receiving haemodialysis. 2022.

32.	Thompson MG, Natarajan K, Irving SA, et al. Effectiveness of a Third Dose of mRNA Vaccines Against COVID-19-Associated Emergency Department and Urgent Care Encounters and Hospitalizations Among Adults During Periods of Delta and Omicron Variant Predominance - VISION Network, 10 States, August 2021-January 2022. *MMWR Morb Mortal Wkly Rep* 2022; **71**(4): 139-45.

**Table S1 Model parameters for cost-effectiveness analysis of COVID-19 booster vaccination in Australia.**

| **Model parameters** | **Values** | **Reference** |
| --- | --- | --- |
| **Epidemiological parameters** |  |  |
| Duration of clinical disease stages of COVID-19 (days) |  | ^17-19^ |
| Incubation | 5.2 (4.1, 7.0) |  |
| Asymptomatic infection | 6.0 (2.0, 12.0) |  |
| Mild/Moderate illness | 10.0 |  |
| Severe illness |  |  |
| In mild/moderate state | 6.5 |  |
| In severe state | 10.5 |  |
| Critical (recover) illness |  |  |
| In mild/moderate state | 3.0 |  |
| In severe state | 7.1 |  |
| In critical state | 11.9 |  |
| In recuperable state | 5.7 |  |
| Critical (die) illness |  |  |
| In mild/moderate state | 3.0 |  |
| In severe state | 7.1 |  |
| In critical state | 11.9 |  |
| **Costing parameters** |  |  |
| Per PCR test for COVID-19 infection, A$ | 85 | ^10^ |
| Per rapid antigen self-test for COVID-19 infection, A$ | 13.25 | ^11^ |
| Per general practitioner (GP) consultation, A$ | 37 | ^12^ |
| Per day of general hospitalization, A$ | 1300.9 | ^13^ |
| Per day of ICU hospitalization, A$ | 4822.3 | ^14^ |
| Per dose of COVID-19 vaccine booster, A$ | 53 | ^15^ |
| Per dose of vaccine administration, A$ | 19.95 | ^16^ |
| Direct medical cost of varied COVID-19 severity, $ |  | ^5,17-19^ |
| Asymptomatic infection | 0 |  |
| Mild/Moderate illness | 37 |  |
| Severe illness | 13696.5 |  |
| Critical (recover) illness | 41046.4 |  |
| Critical (die) illness | 74073.4 |  |
| **Life quality parameters** |  |  |
| Utility weight of clinical disease progression severity |  | ^20-23^ |
| In asymptomatic state | 1 |  |
| In mild/moderate state | 1 |  |
| In severe state | 0.785 (0.700, 0.910) |  |
| In critical state | 0.520 (0.400, 0.730) |  |
| In recuperable state | 0.905 (0.810, 1.000) |  |
| In dead state | 0 |  |
| Discount rate, per year | 3% (0%, 6%) | ^24^ |

**Table S2 Fully vaccinated effectiveness (VE) for SARS-CoV-2 variant (Omicron) infection.**

| No | First author | Duration | Cases  (PCR-positive) | | Control  (PCR-negative) | | Note |
| --- | --- | --- | --- | --- | --- | --- | --- |
|  |  |  | Vaccinated | Unvaccinated | Vaccinated | Unvaccinated |  |
| **Short-term VE for preventing Omicron infection (2 weeks-3 months after 2nd dose)** | | | | | | | |
| ^25^ | Powell FK et al (England, 2022) | Up to Jan 12, 2022 | 115 | 20098 | 379 | 39712 | BNT162b2, ≥14 days |
| ^25^ | Powell FK et al (England, 2022) | Up to Jan 12, 2022 | 4925 | 13517 | 9859 | 20009 | BNT162b2, ≥14 days |
| ^26^ | Collie et al (South Africa, 2022) | Sep 1-Dec 7,2021 | 6290 | 14179 | 26035 | 44477 | BNT162b2, ≥14 days |
| ^2^ | Willett et al (England, 2022) | Dec 22-Dec 28,2021 | 278 | 2179 | 2448 | 15465 | BNT162b2, ≥14 days |
| ^3^ | HHA et al (Qatar, 2022) | Dec 23, 2021-Feb 2, 2022 | 297 | 26258 | 343 | 15922 | BNT162b2, 14-90 days |
| ^27^ | Tseng et al (USA, 2022) | Dec 6-Dec 30,2021 | 245 | 8845 | 836 | 16947 | mRNA-1273, 14-90 days |
| ^2^ | Willett et al (England, 2022) | Dec 22-Dec 28,2021 | 90 | 1991 | 761 | 13778 | mRNA-1273, ≥14 days |
| ^3^ | HHA et al (Qatar, 2022) | Dec 23, 2021-Feb 2, 2022 | 44 | 12841 | 46 | 7754 | mRNA-1273, 14-120 days |
| ^28^ | Buchan et al (Canada, 2022) | Dec 6-Dec 26,2021 | 1234 | 2024 | 9023 | 13704 | BNT162b2&mRNA-1273, 7-120 days |
| ^2^ | Willett et al (England, 2022) | Dec 22-Dec 28,2021 | 45 | 1946 | 468 | 13485 | AZD1222, ≥14 days |
| **Long-term VE for preventing Omicron infection (>3 months after 2nd dose)** | | | | | | | |
| ^27^ | Tseng et al (USA, 2022) | Dec 6-Dec 30,2021 | 10550 | 19150 | 21843 | 37954 | mRNA-1273, ≥90 days |
| ^3^ | HHA et al (Qatar, 2022) | Dec 23, 2021-Feb 2, 2022 | 21205 | 151793 | 12505 | 91090 | BNT162b2, ≥90 days |
| ^3^ | HHA et al (Qatar, 2022) | Dec 23, 2021-Feb 2, 2022 | 8167 | 34284 | 4840 | 20517 | mRNA-1273, ≥120 days |
| ^28^ | Buchan et al (Canada, 2022) | Dec 6-Dec 26,2021 | 12579 | 13369 | 82282 | 86963 | BNT162b2&mRNA-1273, ≥120 days |
| ^27^ | Tseng et al (USA, 2022) | Dec 6-Dec 30,2021 | 10550 | 19150 | 21843 | 37954 | mRNA-1273, ≥90 days |
| **Short-term VE for preventing severe COVID-19 disease (2 weeks-3 months after 2nd dose)** | | | | | | | |
| ^29^ | Tartof et al (USA, 2022) | Dec 1, 2021-Jan 11, 2022 | 49 | 830 | 557 | 3762 | BNT162b2, 7-90 days |
| ^1^ | Ferdinands et al (USA, 2022) | Aug 26, 2021-Jan 22, 2022 | 942 | 14933 | 1595 | 13403 | BNT162b2&mRNA-1273, 14-120 days |
| ^26^ | Collie et al (South Africa, 2022) | Sep 1-Dec 7,2021 | 121 | 341 | 32204 | 58315 | BNT162b2, ≥14 days |
| ^29^ | Tartof et al (USA, 2022) | Dec 1, 2021-Jan 11, 2022 | 93 | 314 | 4421 | 7626 | BNT162b2, ≥7 days |
| ^27^ | Tseng et al (USA, 2022) | Dec 6-Dec 30,2021 | 7 | 21 | 28 | 42 | mRNA-1273, ≥14 days |
| ^30^ | Lauring et al (USA, 2022) | Dec 26, 2021-Jan 14, 2022 | 211 | 479 | 158 | 247 | BNT162b2&mRNA-1273, ≥14 days |
| ^1^ | Ferdinands et al (USA, 2022) | Aug 26, 2021-Jan 22, 2022 | 91 | 1981 | 291 | 2312 | BNT162b2&mRNA-1273, 14-120 days |
| **Long-term VE for preventing severe COVID-19 disease (>3 months after 2nd dose)** | | | | | | | |
| ^29^ | Tartof et al (USA, 2022) | Dec 1, 2021-Jan 11, 2022 | 540 | 1321 | 3864 | 7069 | BNT162b2, ≥90 days |
| ^1^ | Ferdinands et al (USA, 2022) | Aug 26, 2021-Jan 22, 2022 | 7409 | 21400 | 9876 | 21684 | BNT162b2&mRNA-1273, ≥120 days |
| ^1^ | Ferdinands et al (USA, 2022) | Aug 26, 2021-Jan 22, 2022 | 888 | 2778 | 2349 | 4370 | BNT162b2&mRNA-1273, ≥120 days |

**Table S3 Booster vaccination effectiveness (VE) for SARS-CoV-2 variant (Omicron) infection.**

| No | First author | Duration | Cases  (PCR-positive) | | Control  (PCR-negative) | | Note |
| --- | --- | --- | --- | --- | --- | --- | --- |
|  |  |  | Vaccinated | Unvaccinated | Vaccinated | Unvaccinated |  |
| **Short-term booster VE for preventing Omicron infection (2 weeks-3 months after 2nd dose)** | | | | | | | |
| ^28^ | Buchan et al (Canada, 2022) | 2021.12.6-2021.12.26 | 600 | 1390 | 10273 | 14954 | BNT162b2&mRNA-1273+BNT162b2 |
| ^28^ | Buchan et al (Canada, 2022) | 2021.12.6-2021.12.26 | 80 | 870 | 1865 | 6546 | BNT162b2&mRNA-1273+mRNA-1273 |
| ^27^ | Tseng et al (USA, 2022) | 2021.12.6-2021.12.31 | 2464 | 10943 | 9677 | 21602 | mRNA-1273+mRNA-1273 |
| ^29^ | Tartof et al (USA, 2022) | 2021.12.1-2022.1.11 | 152 | 933 | 3240 | 6445 | BNT162b2+BNT162b2 |
| ^31^ | Spensley et al (England, 2022) | 2021.12.1-2022.1.16 | 76 | 91 | 671 | 726 | BNT162b2&AZD1222 +BNT162b2 |
| ^2^ | Willet et al (Scotland, 2022) | 2021.12.22-2021.12.28 | 655 | 2556 | 11581 | 24598 | BNT162b2+mRNA-1273 |
| ^2^ | Willet et al (Scotland, 2022) | 2021.12.22-2021.12.28 | 1193 | 3094 | 21704 | 34721 | BNT162b2+BNT162b2 |
| ^2^ | Willet et al (Scotland, 2022) | 2021.12.22-2021.12.28 | 19 | 1920 | 235 | 13252 | mRNA-1273+BNT162b2 |
| ^2^ | Willet et al (Scotland, 2022) | 2021.12.22-2021.12.28 | 25 | 1926 | 296 | 13313 | mRNA-1273+mRNA-1273 |
| ^2^ | Willet et al (Scotland, 2022) | 2021.12.22-2021.12.28 | 743 | 2644 | 15159 | 28176 | AZD+mRNA-1273 |
| ^2^ | Willet et al (Scotland, 2022) | 2021.12.22-2021.12.28 | 1130 | 3031 | 23641 | 36658 | AZD+BNT162b2 |
| ^3^ | Chemaitelly et al (Qatar, 2022) | 2021.12.23-2022.2.2 | 497 | 51829 | 508 | 31189 | mRNA-1273+mRNA-1273 |
| ^3^ | Chemaitelly et al (Qatar, 2022) | 2021.12.23-2022.2.2 | 3476 | 82424 | 3441 | 49724 | BNT162b2+BNT162b2 |
| **Long-term booster VE for preventing Omicron infection (>3 months after 2nd dose)** | | | | | | | |
| ^29^ | Tartof et al (USA, 2022) | 2021.12.1-2022.1.11 | 21 | 802 | 219 | 3424 | BNT162b2+BNT162b2 |
| ^3^ | Chemaitelly et al (Qatar, 2022) | 2021.12.23-2022.2.2 | 719 | 13858 | 618 | 8429 | BNT162b2+BNT162b2 |
| **Short-term booster VE for preventing severe COVID-19 disease (2 weeks-3 months after 2nd dose)** | | | | | | | |
| ^27^ | Tseng et al (USA, 2022) | 2021.12.6-2021.12.31 | 4 | 18 | 26 | 36 | mRNA-1273+mRNA-1273 |
| ^32^ | Thompson et al (USA, 2022) | 2021.8-2022.1 | 520 | 3918 | 3356 | 6954 | BNT162b2&mRNA-1273+BNT162b2&mRNA-1273 |
| ^32^ | Thompson et al (USA, 2022) | 2021.8-2022.1 | 24 | 198 | 490 | 776 | BNT162b2&mRNA-1273+BNT162b2&mRNA-1273 |
| ^30^ | Lauring et al (USA, 2022) | 2021.12.26-2022.1.14 | 80 | 348 | 135 | 224 | BNT162b2&mRNA-1273+BNT162b2&mRNA-1273 |
| ^3^ | Chemaitelly et al (Qatar, 2022) | 2021.12.23-2022.2.2 | 19 | 259 | 339 | 717 | BNT162b2+BNT162b2 |
| ^3^ | Chemaitelly et al (Qatar, 2022) | 2021.12.23-2022.2.2 | 1 | 206 | 24 | 572 | mRNA-1273+mRNA-1273 |
| ^1^ | Ferdinands et al (USA, 2022) | 2021.8-2022.2 | 1696 | 15687 | 8514 | 19322 | BNT162b2&mRNA-1273+BNT162b2&mRNA-1273 |
| ^1^ | Ferdinands et al (USA, 2022) | 2021.8-2022.2 | 240 | 2130 | 2404 | 4425 | BNT162b2&mRNA-1273+BNT162b2&mRNA-1273 |
| **Long-term booster VE for preventing severe COVID-19 disease (>3 months after 2nd dose)** | | | | | | | |
| ^1^ | Ferdinands et al (USA, 2022) | 2021.8-2022.2 | 242 | 14233 | 721 | 11529 | BNT162b2&mRNA-1273+BNT162b2&mRNA-1273 |
| ^1^ | Ferdinands et al (USA, 2022) | 2021.8-2022.2 | 36 | 1926 | 153 | 2174 | BNT162b2&mRNA-1273+BNT162b2&mRNA-1273 |
